# A Systematic Review of Droplet and Aerosol Generation in Dentistry

**DOI:** 10.1101/2020.08.28.20183475

**Authors:** N. Innes, I.G. Johnson, W. Al-Yaseen, R. Harris, R. Jones, S. Kc, S. McGregor, M. Robertson, W.G. Wade, J.E. Gallagher

**Author notes:** **Corresponding author: Prof Nicola Innes** Professor Nicola P T Innes, Head of School of Dentistry, Professor and Honorary Consultant, Paediatric Dentistry, College of Biomedical & Life Sciences, Cardiff University, Heath Park, Cardiff, CF14 4XY, Web: https://www.cardiff.ac.uk/dentistry.

## Abstract

**Objectives:** Against the COVID-19 pandemic backdrop and potential disease transmission risk by dental procedures that can generate aerosol and droplets, this review aimed to identify which clinical dental procedures do generate droplets and aerosols with subsequent contamination, and for these, characterise their pattern, spread and settle.

**Data Sources:** Six databases were searched and citation chasing undertaken (to 11/08/20).

**Study selection:** Screening stages were undertaken in duplicate, independently, by two researchers. Data extraction was performed by one reviewer and verified by another.

**Results:** Eighty-three studies met the inclusion criteria and covered: Ultrasonic scaling (USS, n=44), highspeed air-rotor (HSAR, n=31); oral surgery (n=11), slow-speed handpiece (n=4); air-water (triple) syringe (n=4), air-polishing (n=4), prophylaxis (n=2) and hand-scaling (n=2). Although no studies investigated respiratory viruses, those on bacteria, blood splatter and aerosol showed activities using powered devices produced the greatest contamination. Contamination was found for all activities, and at the furthest points studied. The operator’s torso operator’s arm, and patient’s body were especially affected. Heterogeneity precluded significant inter-study comparisons but intra-study comparisons allowed construction of a proposed hierarchy of procedure contamination risk: higher risk (USS, HSAR, air-water syringe [air only or air/water together], air polishing, extractions using motorised handpieces); moderate (slow-speed handpieces, prophylaxis with pumice, extractions) and lower (air-water syringe [water only] and hand scaling.

**Conclusion:** Significant gaps in the evidence, low sensitivity of measures and variable quality limit firm conclusions around contamination for different procedures. However, a hierarchy of contamination from procedures can be proposed for challenge/verification by future research which should consider standardised methodologies to facilitate research synthesis.

**Clinical significance:** This manuscript addresses uncertainty around aerosol generating procedures (AGPs) in dentistry. Findings indicate a continuum of procedure-related aerosol generation rather than the current binary AGP or non-AGP perspective. This informs discussion around AGPs and direct future research to help support knowledge and decision making around COVID-19 and dental procedures.

## Background

SARS-CoV-2 is the highly infectious virus which causes COVID-19 [1]. Transmission has been thought to be primarily via respiratory droplets, direct contact and fomites. Virus particles can be detected up to 72 hours after inoculation of plastic and steel surfaces [1-3]. However, SARS-CoV-2 has been found to remain viable in air for at least three hours with growing evidence and concern over possible airborne transmission [2, 4].

Dental care involves close patient contact for prolonged periods leading to concern over transmission through aerosol generation during dental procedures [5, 6], compounded by consistent detection of SARS-CoV-2 in saliva [7]. In dentistry, universal precautions have been standard practice, based on evidence-informed infection control. These evolve as evidence emerges, particularly in response to blood and water borne infections and prion transmission [8-10].

The term Aerosol Generating Procedure (AGP) has been described as, “any procedure on a patient that can induce the production of aerosols of various sizes” although there is currently no agreed definition and a confusing lack of consistency in terminology. The UK National Emerging Respiratory Virus Threats Advisory Group has described “dental procedures (using highspeed devices such as Ultrasonic scalers and highspeed drills)” [11] as posing an increased risk of respiratory infection transmission. New terms such as aerosol generating exposure (AGE) have been suggested [12]. Policy documents have focused on ultrasonic scalers (USS), high-speed air-rotors (HSAR), air-water syringes (also known as triple or 3-in-1), and air polishers as sources of aerosols, with rubber dam and high-volume suction as mitigating measures [13-15].

To manage transmission risk of SARS-CoV-2, the extent and contamination of droplets and aerosols involved in dental procedures need to be identified. Similarly, the pattern, and timing, associated with spread and settle of droplets and aerosols in the context of clinical dentistry need to be understood to inform policy on surgery fallow times between patients. Globally specified patient spacing times for AGPs vary from none to 120 minutes [16].

An aerosol is defined as a suspension of liquid or solid in air [17, 18]. When an aerosol is created with a liquid, a wide range of droplet sizes are produced. Particle size is a continuum, from larger heavier droplets, > 5 µm in diameter that fall rapidly to the ground, typically within 1 m of the source as splatter. Aerosols are composed of droplet nuclei ≤ 5 µm in diameter and can remain suspended in air for many hours and be moved by air currents. At present, dental procedures are categorised dichotomously as either aerosol producing or non-aerosol producing. The former refers to procedures considered to produce smaller droplets of ≤ 5 µm and the latter referring to procedures that are considered to produce few or no smaller droplets but may still produce larger droplets (> 5 µm). For the purposes of this review, aerosol will refer to suspensions of particles ≤ 5 µm in diameter.

This review aims to critically assess existing knowledge and reduce uncertainty around dental procedures that generate droplets and aerosol, supporting policy making and local IPC protocols.

## Research question

What is known and what is not known about aerosols and droplets relevant to clinical dentistry?

Objectives:

1. To identify and catalogue activities within clinical dentistry and the dental surgery that generate aerosols and droplets
2. For these activities, to:

a. Characterise the pattern of droplet and aerosol spread and settle relevant to the dental surgery and dental laboratories
b. Identify whether there is evidence of an association with exposure, infection and transmission of pathogenic micro-organisms
c. List micro-organisms that have been studied
d. Record outcomes and outcome measures
3. To identify gaps in the evidence related to aerosols and droplets relevant to clinical dentistry

## Methods

### Protocol and Registration

This review has been conducted and reported according to the Preferred Reporting Items for Systematic Reviews and Meta-Analyses (PRISMA) [19], registered under the International Prospective Register of Systematic Reviews ID number CRD42020193058 and Appendix 1 gives full details.

### Eligibility Criteria for study selection

Inclusion criteria:

- Study methodology – including but not limited to; trials, observational, experimental (including those using manikins, modelling studies, etc.), qualitative studies, non-clinical reports and other relevant studies;
- Topic of study -investigate activities that generate aerosols etc. relevant to clinical dentistry;
- Where there is a measure of aerosols and droplets;
- Types of settings: dental practices and hospital settings, including simulated environments where there are relevant to the conduct of dental procedures and investigations; and
- English or Chinese language

### Information Sources

Medline (OVID), Embase (OVID), Cochrane Central Register of Controlled Trials, Scopus, Web of Science and LILACS databases were searched for studies meeting the inclusion criteria and ClinicalTrials.gov was searched for recently completed, ongoing, or recruiting trials from the start of the databases to May 2020. The search was updated on 11 August 2020 to identify new studies published since the original search was conducted.

### Search

The search strategy (Appendix 2) comprised controlled vocabulary and keywords. The references of all reviews, policy documents and included studies were screened for eligible studies.

### Screening and selection of studies

Titles and abstracts were deduplicated and screened in Rayyan [20] independently and in duplicate by two reviewers. Where either reviewer considered a paper potentially eligible for inclusion, the full text was sought. Full texts of potential articles were retrieved and assessed independently and in duplicate. Full texts were exported into Endnote and a database created in Excel. Differences were resolved by consensus involving at least one other research group member.

### Data extraction

A standardised data extraction form was developed a priori and refined based on repeat pilot testing with a minimum of five publications and three data extractors. Eight reviewers were trained in data extraction form completion. Reviewers extracted data into an excel spreadsheet singly but consulted another reviewer where data reporting was unclear. Key missing data items were managed by contacting study investigators where possible. For studies where an intervention was measured for its ability to alter droplet and aerosol spread, only data relating to the baseline or control (i.e. without the intervention effect) was extracted.

### Data items

The items of data extracted included: study demographics; dental procedures investigated; methodology; findings – (related to the reviews’ outcomes). Detection methods for contamination were categorised as microbial, blood and other (non-microbial/non-blood) methodologies.

### Quality assessments

The quality of the papers (see Protocol in Appendix 1) was assessed. Increases in evidence production aligning to infectious disease outbreaks was checked to investigate publication bias.

### Sensitivity assessments

The sensitivity of contamination assessment tools was assessed using schema tailored to the individual methodologies: microbial measures; blood measures; other [non-microbial/non-blood measures). These are presented overall for all studies and grouped by procedure to allow a picture of the sensitivity of the methods used, and therefore accuracy of the results in reflecting actual contamination. This allowed a judgement on the likelihood of under-or over-reporting of contamination for each study, and by procedure.

### Relative contamination of procedures

Where methodology was similar enough to compare contamination levels or studies included multiple procedures, the relative contamination levels between procedures were examined.

### Results

There were 83 studies (Appendix 3) which met the inclusion criteria and for which we could obtain full manuscripts (see PRISMA flow chart Figure 1.).

**Figure 1.**
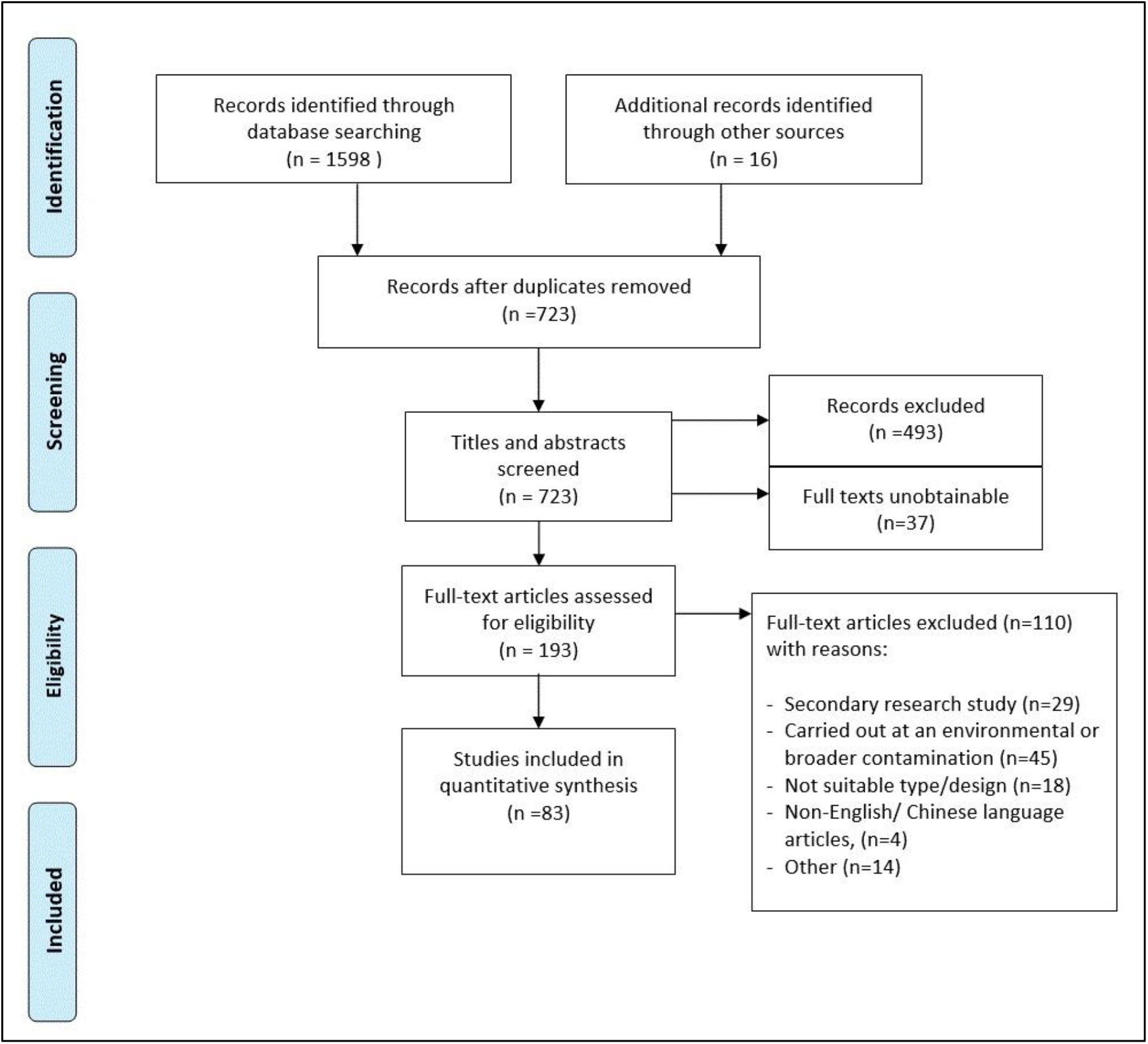
Preferred Reporting Items for Systematic Reviews and Meta-Analyses (PRISMA) (Moher et al. 2009) flow chart. A full description of study characteristics can be found in Appendix 4.

Studies originated from 24 countries, the majority conducted in the USA (n=26) and India (n= 21). They were published between 1963 to 2020 with 43/83 (52%) of the studies published in the last decade (Figure 2.)

**Figure 2.**
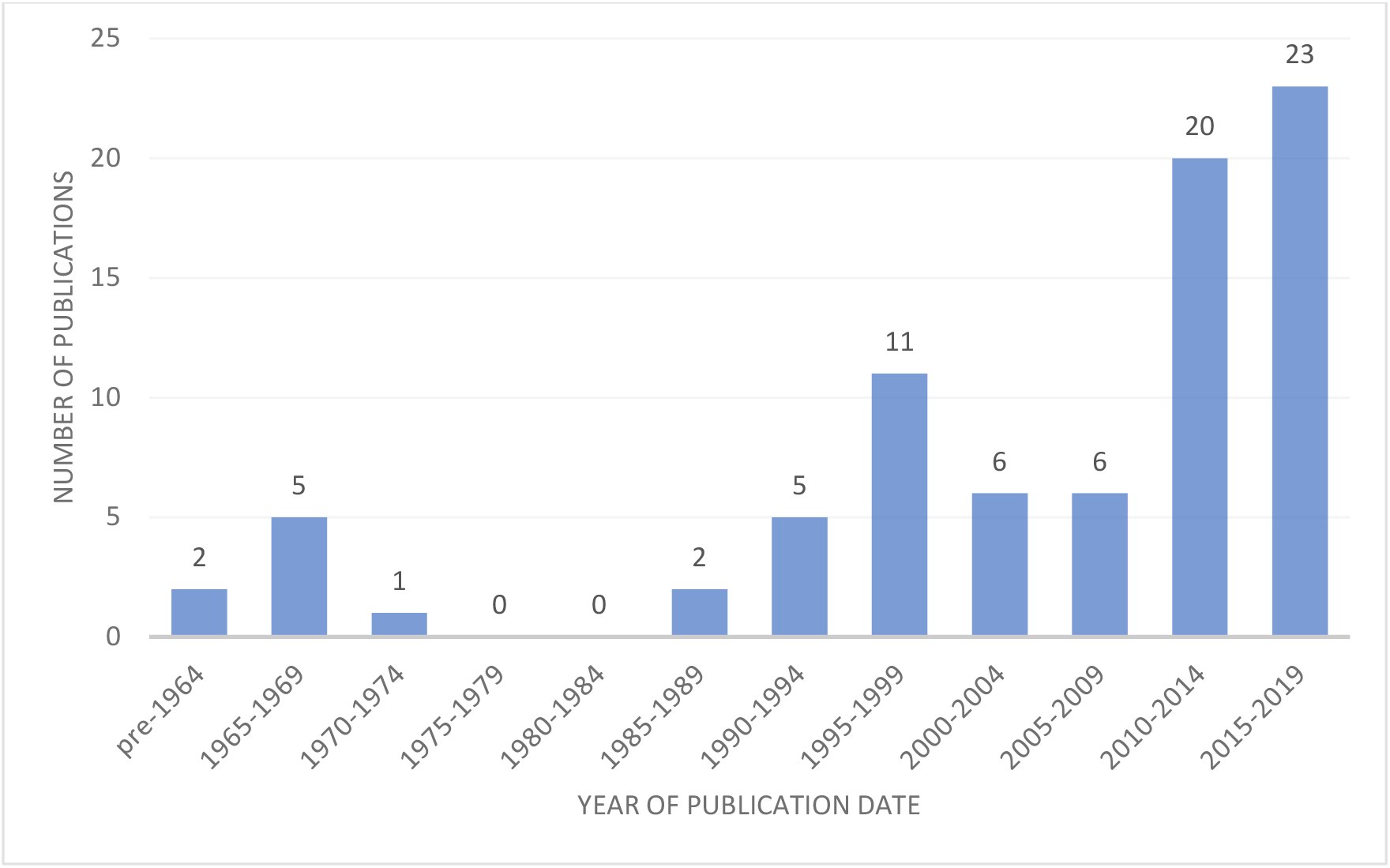
Publications by date (n=81; 2 publications from 2020 were not included).

A full description of the studies’ characteristics and the data extracted can be found in Appendix 4.

The extracted data were heterogeneous across key characteristics, including; aims, methodology and outcomes. A narrative summary was undertaken and within study comparisons of relative contamination made. Whilst there were no studies showing a direct association between dental procedures with exposure, infection and transmission of pathogenic micro-organisms (outcome 2b), there was evidence of contamination of persons in the dental surgery and the surgery environment (surfaces, equipment etc.) and air from all procedures investigated although the levels of contamination varied (Appendix 4).

Approaches to the investigation varied: some studied procedures, some instruments and some both. Data were separated into the following categories: USS (n=44 studies), HSAR (n=31); oral surgery (n=11), slow-speed handpiece (n=4); air-water syringe (n=4), air polishing (n=4), prophylaxis with cup and pumice (n=2) and hand scaling (n=2).

**Table 1.** Main characteristics of the included studies categorised by dental procedure/ instrument use (n=83). Please note that the numbering in this table refers to the list of references (included studies) in Appendix 3 and are not the same numbering system as found in the main text.

| Procedure category (no of studies)<br>(First author year) Unique study identification numbers | Procedures investigated (e.g. cavity preparation) and type of investigation (clinical/ laboratory, patient/ mannequin) | Investigated contamination of operator, assistant, patient | Area within dental surgery investigated* | Different timepoints sampled (hours, minutes, seconds) * Unique study identification numbers | Sensitivity scores for outcome methodologies |
| --- | --- | --- | --- | --- | --- |
| **USS (n=44)**<br>Balcos 2019<sup>5</sup>, Barnes 1998<sup>6</sup>, Bentley 1994<sup>8</sup>, Choi 2018<sup>9</sup>, Chuang 2014<sup>10</sup>, Devker 2012<sup>16</sup>, Feres 2010<sup>21</sup>, Fine 1992<sup>22</sup>, Fine 1993 a<sup>23</sup>, Fine 1993b<sup>24</sup>, Graetz 2014<sup>25</sup>, Grenier 1995<sup>29</sup>, Gupta 2014<sup>31</sup>, Hallier 2010<sup>32</sup>, Harrel 1998<sup>34</sup>, Harrel 1996<sup>33</sup>, Holloman 2015<sup>37</sup>, Jawade 2016<sup>41</sup>, Kau r 2014<sup>44</sup>, King 1997<sup>45</sup>, Labaf 2011<sup>48</sup>, Miller 1971<sup>90</sup>, Mohan 2016<sup>52</sup>, Narayana 2016<sup>54</sup>, Nejatidanesh 2013<sup>55</sup>, Prospero 2003<sup>59</sup>, Purohit 2009<sup>60</sup>, Ramesh 2015<sup>61</sup>, Rao 2015<sup>62</sup>, Reddy 2012<sup>64</sup>, Retamal-Valdes 2017<sup>65</sup>, Rivera-Hidalho 1999<sup>66</sup>, Sadun 2020<sup>69</sup>, Saini 2015<sup>70</sup>, Sawhney 2015<sup>71</sup>, Serban 2013<sup>72</sup>, Sethi 2019<sup>73</sup>, Shetty 2013<sup>74</sup>, Singh 2016<sup>76</sup>, Swaminathan 2014<sup>78</sup>, Timmerman 2004<sup>80</sup>, Veena 2015<sup>83</sup>, Watanabe 2013<sup>85</sup>, Yamada 2011<sup>87</sup> | Ultrasonic scaling (n=44)<br>Simulation Mannequin (n=3)<sup>8,25,83</sup><br>Simulation Typodont Laboratory (n=3)<sup>33,34,66</sup> | **Operator (n=18)**<br>5,8,9,10,16,21,31,44,45,55, 59,60,63, 65,70,72,83,85<br>Head/face (n=13)<sup>8,9,10,16,21,31,44,53,59,65,72,83,85</sup><br>Body (n=8)<sup>5,8,10,31,60,70,83,85</sup><br>**Assistant (n=4)**<br>Head (n=2)<sup>16,83</sup><br>Body (n=3)<sup>31,70,83</sup><br>**Patient (n=14)**<br>Chest (n=14)<sup>5,8,16,21,31,44,60,61,62,65,70,71,73,85</sup> | Environmental surfaces < 1m (n=15)<br>8,16,21,33,37,45,52,54,61,66,71,73,74,76,78<br>>1m (n=3)<br>25,44,64<br>< 1m and > 1m (n=9)<br>10,41,48,69,90,70,80,83,87<br>Air Samples (n=7)<sup>6,22,23,24,29,32,87</sup> | During procedure (n=28)<br>8,9,22,23,24,25,32,48,90,52,54,59,60,61,62,64,65,66,69,70,72,73,74,76,78,80,85,87<br>During and after combined (n=8)<br>10,16,21,31,41,44,45,71<br>After procedure (n=3)<br>0-30 min<sup>29,37,83</sup><br>30 min-1 hr<sup>29,37,83</sup><br>1-2 hr<sup>29,83</sup><br>2-4 hr<sup>29</sup> | **High sensitivity** (n=5)<br>**Moderate sensitivity** (n=3)<br>**Low sensitivity** (n=34)<br>**Uncategorized sensitivity** (n=2) |
| **HSAR (n= 31)**<br>Al-Amad 2017<sup>3</sup>, Belting 1963<sup>93</sup>, Bentley 1994<sup>8</sup>, Cochrane 1989<sup>92</sup>, Dahlke 2012<sup>13</sup>, Day 2008<sup>15</sup>, Earnest<sup>20</sup>, Greco 2008<sup>28</sup>, Grenier 1995<sup>29</sup>, Grundy 1967<sup>30</sup>, Hallier 2010<sup>32</sup>, Hausler 1966<sup>36</sup>, Junevičius 2005<sup>42</sup>, Labaf 2011<sup>48</sup>, Larato 1966<sup>91</sup>, Manarte-Monteiro 2013<sup>50</sup>, Micik 1969<sup>89</sup>, Miller 1971<sup>90</sup>, Miller 1995<sup>51</sup>, Neiatidanesh 2013<sup>55</sup>, Oliveira 2018<sup>56</sup>, Prospero 2003<sup>59</sup>, Purohit 2009<sup>60</sup>, Rautemaa 2006<sup>63</sup>, Samaranayake 1989<sup>88</sup>, Stevens 1963<sup>96</sup>, Tag El-Din 1997<sup>94</sup>, Toroğlu 2001<sup>81</sup>, Toroğlu 2003<sup>8</sup>, Travaglini<sup>95</sup>, Yamada 2011<sup>87</sup> | Tooth/cavity preparation (clinic) (n= 24)<br>Mannequin/ Typodont (clinic) (n=3)<sup>8,36,42</sup><br>Mannequin/ typodont simulation (laboratory) (n=4)<sup>13,30,51,89</sup> | **Operator (n=10)**<br>Head/face (n=9)<sup>96,95</sup><br>20,8,81,59,63,55,3<br>Body (n=2)<sup>8,60</sup><br>**Assistant (n=3)**<br>Head/face (n=3)<sup>8,81,63</sup><br>Body (n=1)<sup>8</sup><br>**Patient (n=6)**<br>Intra-oral (n=1)<sup>20</sup><br>Head/face (n=1)<sup>95</sup><br>Body (n=4)<sup>92,20,8,94</sup> | Using settle plate within 1m of operating area (n=12)<sup>93,8,92,42,48,59,63,90,60,81,94,87</sup><br>Using settle plate 1m or more from operating area (n=9)<sup>93,48,90,56,63,94,60,63,81</sup> | Within the first 30 minutes after the procedure (n=4)<sup>8,29,51,94</sup><br>Between (30 min -1 hr) after procedure (n=2)<sup>29,51</sup><br>1-2 hr (n=2)<sup>29,51</sup><br>2-4 hr (n=2)<sup>29,51</sup><br>> 4 hr (n=1)<sup>51</sup> | **High sensitivity** (n=4)<br>**Moderate sensitivity** (n=6)<br>**Low sensitivity** (n=21) |
| **Oral surgery (including extractions) (n=11)**<br>Al-Eid 2018<sup>4</sup>, Divya 2019<sup>17</sup>, Hallier 2010<sup>32</sup>, Ishiharma 2008<sup>38</sup>, Ishiharma 2009<sup>39</sup>, Janani 2018<sup>40</sup>, Jimson 2015<sup>43</sup>, Kobza 2018<sup>46</sup>, Wada 2010<sup>84</sup>, Yamada 2011<sup>87</sup>, Aguilar-Duran 2020<sup>98</sup> | Surgical removal of teeth (clinic) (n=8)<sup>4, 17, 38, 39, 84, 43, 87, 98</sup><br>Implants (clinic) (n=1)<sup>98</sup><br>Extraction (clinic) (n=2)<sup>32, 98</sup><br>Other: restorative and periodontal procedures (clinic) (n=1)<sup>87</sup><br>Not verified/ unclear (clinic) (n=2)<sup>40, 46</sup> | **Operator (n=5)**<br>Head/ face/neck (n=4)<sup>4, 38, 40, 98</sup><br>Arms/ Cuffs/ hands (n=3)<sup>4, 38, 40</sup><br>Abdomen/ shoes (n=3)<sup>4, 38, 40</sup><br>Nearby operator (n=1)<sup>43</sup><br>**Assistant (n=3)**<br>Head/ face/neck (n=2)<sup>4, 98</sup><br>Arms/ Cuffs/ hands, (n=1)<sup>4</sup><br>Other body parts (n=1)<sup>4</sup><br>Nearby assistant (n=1)<sup>43</sup><br>**Patient: (n=3)**<br>Head/ face, (n=1)<sup>4</sup><br>Chest (n=2)<sup>4, 17, 43</sup> | Environment and surfaces within 1m from operating area (n=4)<sup>17, 32, 39, 46, 87</sup><br>Environment and surfaces 1m or more from operating area (n=0)<br>Sampled at mouth (n=0) | Up to 30 min during procedure (n=2)<sup>17, 43</sup><br>During treatment with agar plates replaced every 10 min (n=1)<sup>32</sup> | **High** sensitivity (n=3)<br>**Moderate** sensitivity (n=2)<br>**Low** sensitivity (n=6) |
| **Slow-speed handpiece (n=5)**<br>Agostinho 2004<sup>2</sup>, Dawson 2016<sup>14</sup>, Day 2008<sup>15</sup>, Ireland 2003<sup>97</sup>, Kritivasan 2019<sup>47</sup> | Removal of excess material following fixed orthodontic appliance removal (n=3)<sup>14, 15, 97</sup><br>Polishing and trimming of denture (n=2)<sup>2, 47</sup> | **Operator (n=1)<sup>2</sup>**<br>(thorax and abdomen area of technician) | Air sampler 30cm from patient's mouth (n=2)<sup>14, 15</sup><br>Air sampler 10cm from patient's mouth (n=1)<sup>97</sup><br>Settle plates at 1ft, 2ft and 3ft from the motor n=1<sup>47</sup> | Not specified (n=5) | **High** sensitivity (n=3)<br>**Moderate** sensitivity (n=0)<br>**Low** sensitivity (n=2) |
| **Air-water syringe (n=4)**<br>**Belting 1963<sup>93</sup>, Micik 1969<sup>89</sup>, Miller 1971<sup>90</sup>, Miller 1995<sup>51</sup>** | Air-water syringe with air and water used together (n=3)<sup>89, 90, 95</sup><br>Air-water syringe with air alone (n=4)<sup>51, 89, 90, 93</sup><br>Air-water syringe with water alone (n=4)<sup>51, 89, 90, 93</sup> | N/A | Experimental simulation-Air Samples<br>Human aerosol test chamber<sup>89</sup><br>Quartz crystal microbalance cascade impactor (QCMCI)<sup>95</sup><br>Clinical (splatter plate)<sup>90, 91</sup><br>0-1m<sup>90, 91</sup><br>1-2m<sup>90, 91</sup> | During procedure (n=3)<sup>89, 90, 93</sup><br>After procedure (n=1)<sup>51</sup>: at 2 min, 35 min, 2 hr, 4 hr and 6 hr. | **High** sensitivity (n=0)<br>**Moderate** sensitivity (n=1)<br>**Low** sensitivity (n=3) |
| **Air polishing (n=4)** | Air polishing (clinic) | **Operator**<br>Head/face | Air sampled 12 in from | 30 min including procedure of 2 | **High** sensitivity (n=0) |
| Dos Santos 2014<sup>18</sup>, Harrel 1999<sup>35</sup>, Logothetis 1995<sup>49</sup>, Muzzin 1999<sup>53</sup> | (n=3)<sup>18,49,53</sup><br>Mannequin/typodont simulation (laboratory) (n=1)<sup>35</sup> | (n=3)<sup>18,49,53</sup><br>**Patient**<br>Body (n=1)<sup>18</sup> | patients' mouth at a 50 degree angle<sup>53</sup><br>Air sampled < 1m of operating area<sup>35</sup><br>Surface < 1m operating area (n=1)<sup>49</sup><br>Surface > 1 m (n=1)<sup>49</sup> | min<sup>49,53</sup><br>During procedure (n=2)<sup>18,35</sup> | **Moderate** sensitivity (n=1)<br>**Low** sensitivity (n=3) |
| **Prophy: cup and pumice (n=2)**<br>Micik 1969<sup>89</sup>, Miller 1971<sup>90</sup> | Prophy on patient's head in closed experimental test chamber with side glove ports (clinic) (n=1)<sup>89</sup><br>Prophy (clinic) (n=1)<sup>90</sup> | N/A | Air sampling of the closed chamber around the patient's head<sup>89</sup><br>Air sampled 3 ft from floor and 1 ft from patients' mouth (in front) to the sides and end of the surgery<sup>90</sup> | During the procedure (10-120 sec) with 4 min clearance period of quiet breathing before and after<sup>89</sup><br>During procedure (30 sec)<sup>90</sup> | **High** sensitivity (n=0)<br>**Moderate** sensitivity (n=0)<br>**Low** sensitivity (n=2) |
| **Hand scaling (n=2)**<br>Harrel 1998<sup>34</sup>, Micik 1969<sup>89</sup> | Hand scaling patient head in closed test chamber with side glove ports (clinic) (n=1)<sup>89</sup><br>Mannequin/typodont simulation (laboratory) (n=1)<sup>34</sup> | N/A | Air sampling of the closed chamber around the patient's head<sup>89</sup><br>Air sampled around mouth distance not specified<sup>34</sup> | During procedure (10-120 sec) with 4 mins clearance period of quiet breathing before and afterwards<sup>89</sup><br>During procedure (2 sec)<sup>34</sup> | **High** sensitivity (n=0)<br>**Moderate** sensitivity (n=0)<br>**Low** sensitivity sampling/measuring approach (n=3) |

\*m = meter(s); ft – foot/feet; in = inch(es); cm = centimetre(s); hr = hour(s); min = minute(s); sec = second(s).

Settle plates were used in 48 studies, 12 used visual inspection and 23 used air samplers (specific for aerosol). The main findings for each of the instruments/ procedure categories is summarised below (n= number of studies), with further detail provided in Table 1 and Appendix 4.

### USS (n=44 studies)

All 37 studies with measures for droplet contamination (splatter) and where samples were collected from the air (n=7) had positive findings including those that used high volume and standard suction. There was greater droplet contamination within 1m distance of the patient (n=8) [21-28]. The operator’s face (mask and visor area) were heavily contaminated (n=10) as were areas closer to the patient (operator’s nearest arm) (n=2) [27, 29]. When assessed, contamination was identified on the assistant’s face and arm (n=4). The patient’s body was heavily contaminated and their chest one of the most heavily contaminated areas. Contamination levels reduced with increasing distance from the mouth. At the maximum measured distance of 3 m, contamination was identified. Air sampling (n=3) found contaminated aerosol was generated during treatment [27, 30, 31] with a small proportion detectable at four feet before returning to baseline two hours post-treatment [30].

### HSAR (31 studies)

There was wide variability in the procedures investigated: Restorative (n=27); cavity preparation, gaining endodontic access, fixed prosthesis tooth preparation; and Orthodontic (n=4); cement removal [following fixed appliances]; and procedural time (10 seconds to four hours). Contamination was detected on settle plates on surfaces across all (n=15) areas measured up to three meters from patient mouth [23, 32]. Contamination levels were highest in front of the patient and reduced with increasing distance from the mouth [21, 32, 33] and the lowest areas of contamination levels were behind the patient [33]. One study found approximately 80% of aerosol settled immediately following the procedure and reached baseline levels after two hours [30]. The operator, assistant and patient were consistently contaminated, most heavily on the operator’s head and patient’s chest. One study directly compared both single and multi-surgeries and found that contamination was detected over a larger distance when there were multiple dental chairs in an area [30].

### Oral surgery (n= 11)

Oral surgery involving removal of teeth, generally third molars, used motorised handpieces of variable speeds (n=10; 1 assumed), reporting the use of irrigation (n=6), intra-oral suction/aspiration (n=5) in single and multiple chair dental settings, found risk of contamination (mostly blood, with evidence of anaerobic bacteria). Contamination was present on the patient (chest and face), operator (face/head, arm/glove/cuff, chest, abdomen, leg) and assistant (face/head, arm/glove/cuff, body) as well as the dental operatory and air environment (< 1 meter). Surgeons and assistants wore surgical PPE including gowns and the research was conducted in a range of settings, mainly dental hospital outpatient facilities (n=10). Most evidence was of blood spatter (visible and imperceptible) whilst microbiological examination was limited to aerobic testing. Imperceptible blood splatter was significantly higher than visible stains. Very limited evidence on extractions (n=2) suggests lower risk but not without risk of contamination. Risk increased with time, type of procedure and decreased with distance.

### Slow-speed handpiece (n=5)

Three studied removal of excess material following fixed orthodontic appliance treatment [34-36] with air sampling equipment (to detect aerosol). Findings varied but could be related to study design differences. Dawson et al. [36] and Day et al. [35] found a marked increase in bacterial load during debonding and enamel cleaning compared to baseline levels. Ireland et al. [34] detected particles (2µm to >30µm diameter) demonstrating facilitation of aerosol and droplets.

With denture polishing and trimming [37, 38] microbiological contamination was highest 2 feet from the operatory position compared to 1ft and 3ft [37] and microorganisms such as yeasts and Gram-negative bacteria were in the aerosol generated [38].

### Air-water syringe (n=4)

All studies identified contamination following use [21, 39-41]. However, the extent of contamination varied widely (n=3). Air and water used together (spray) generated more than air alone and water alone the least [21, 39, 40]. Smaller particles remained in the air for more than 6 hours [41]. Bacterial contamination from droplets was detectable at 4 feet from the patient [39] and 6 feet from patients [21]. These were the maximum points sampled

### Air polishing (n=4)

Air polishing demonstrated contamination of, in ascending order, the operator’s forehead, operator’s mouth and patients’ chest. Contamination from air polishing was found nine feet from the treatment area even in a surgery with 13 air changes/hour [42].

### Prophylaxis (n=2)

Prophylaxis with cup and pumice (n=2), [21, 40] produced less contamination than air-polishing (n=4). It produced a higher rate of contamination than washing teeth with a water stream, but lower than drying teeth with an air spray or using a high turbine with water coolant, as measured using the same closed test chamber (n=1).

### Hand scaling (n=3)

Hand scaling produced minimal contamination (two artificial environments; patients’ head in a closed experimental test chamber with side glove ports for the operator (n=1), and a mannequin in a closed box (n=1). Levels of air contamination in the test chamber were comparable to a clinical examination in the same experiment. When hand scaling with orthodontic treatment was compared to HSAR contamination was much lower when no powered instruments were used [33].

### Relative contamination levels

Although there were 83 studies, the degree of heterogeneity in methodology meant it was not possible to compare data between them. The outcome of interest was contamination. It could be grouped as being microbial, blood and non-microbial/non-blood. However, within these, there were a large variety of outcome measures (Appendix 4). For example, even within those looking at colony forming units, the outcome measures encompassed; whole plate (CFU/mm^2^); (CFU/mm^3^); (CFU/cm^2^); (CFU/m^3^), volume of sampled air (CFU/cm^2^ /min), Index of Microbial Air Contamination (CFU/m^2^ /h) and rate of production (CFU/min).

However, it was evident that there was a hierarchy of contamination levels with some procedures generating more contamination than others. A network diagram (Figure 3) illustrates where intra-study comparisons exist. Data from these 13 studies were tabulated (Appendix 5, Table 1) and compared for relative contamination levels within studies (Appendix 5, Table 2).

**Figure 3.**
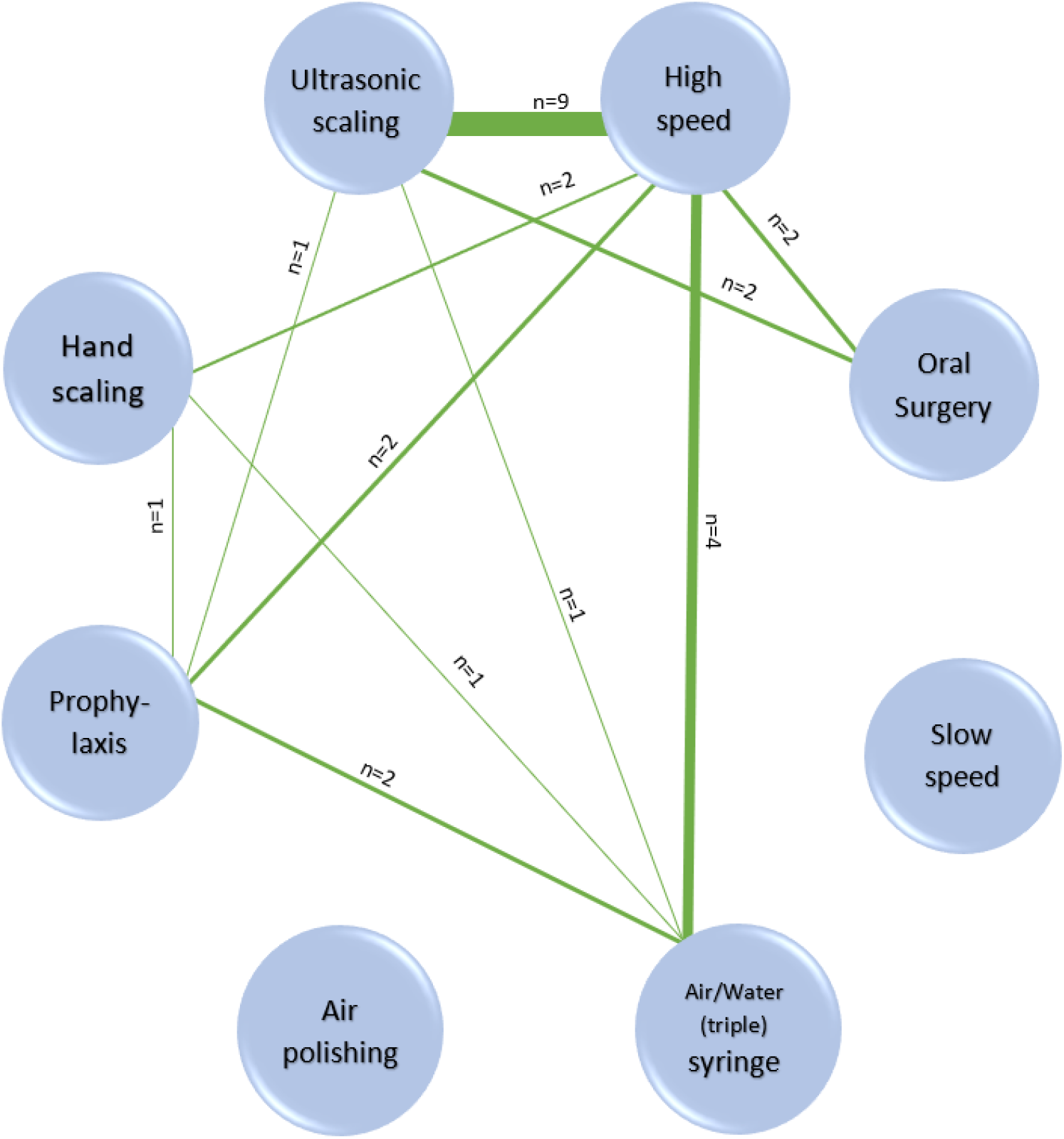
Network diagram illustrating where studies included comparison between different procedures within them. (See also Appendix 5). The nodes represent the eight procedures and the lines between them show where a study compares them. The number of studies is shown by ‘n=’ and also the relative thickness of the lines. Where a node has no linkages, there are no studies comparing it with another procedure.

These studies and their data were then categorised as higher, moderate and lower relative contamination levels (Figure 4). This is a proposed hierarchy model and it should be noted that positions denote relative positions along a spectrum rather than definitive cut-offs between the three levels.

**Figure 4.**
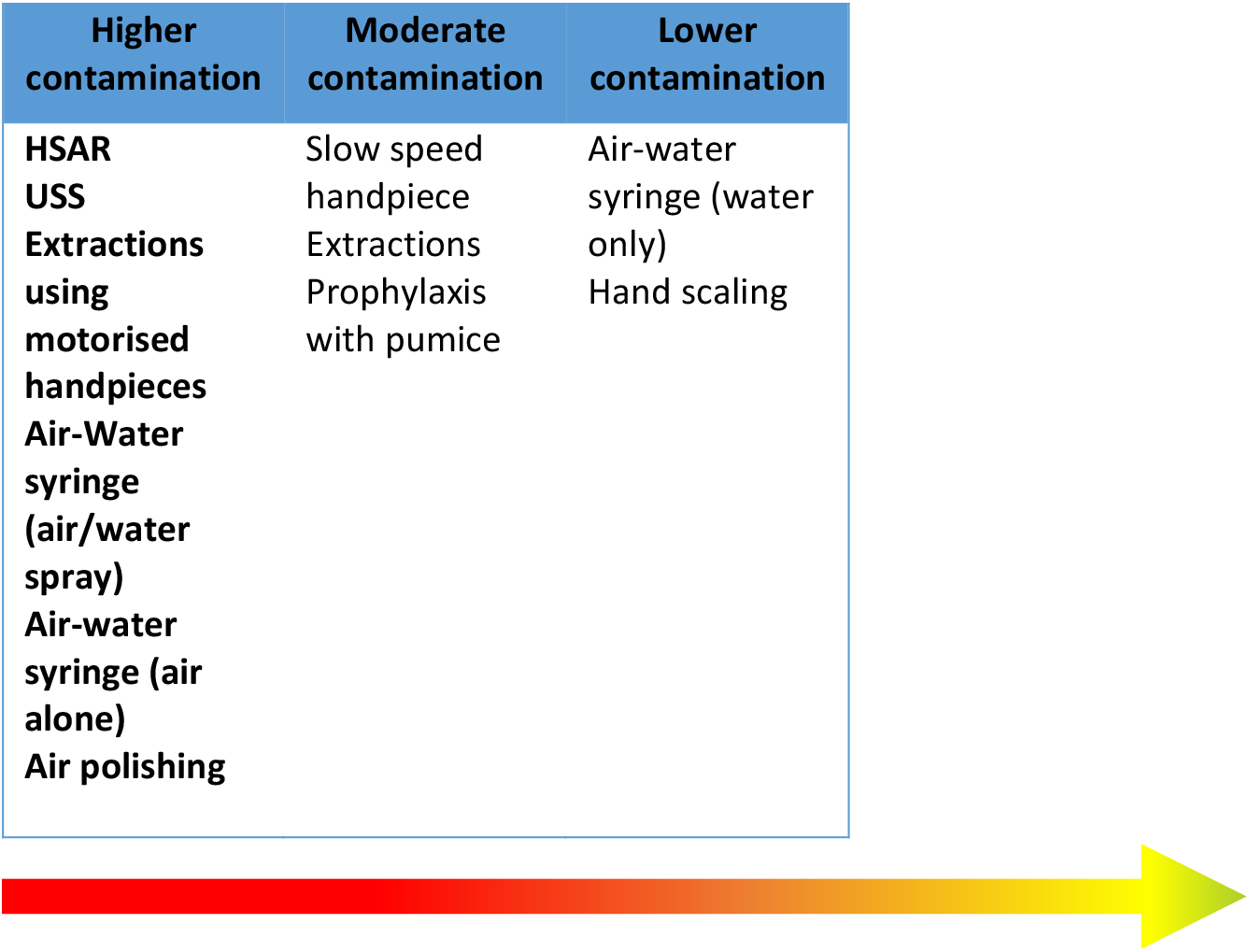
Proposed levels of contamination associated with different procedures, drawn from Appendix 5 showing levels of contamination within studies to minimize dissimilarities in methodology, procedures and outcomes that might account for differences. Note that this must be interpreted with caution and will need to be modified as further evidence becomes available. *Indicates very low certainty.

### Quality Assessment

The quality assessment (Appendix 6) for the studies showed a mixed picture for each of the seven domains; the majority of studies scored “high” quality for one domain (controls), “moderate” for four (study funding, conflict of interest, procedure description and outcome reporting) and “low” for two (equipment use and sample size).

### Sensitivity

Across all studies, for detecting contamination, 59 were rated as low sensitivity, 11 were moderate and 11 as having a high sensitivity. Sensitivity gradings for each procedure and by study are detailed in Appendix 6.

## Discussion

The 83 studies included in this review looked at eight different activities classified as; USSs, HSAR and slow-speed handpieces, oral surgery, air-water syringe, air polishing and hand scaling. There was heterogeneity between methodologies used to investigate contamination and a lack of consistency even when the same methodology was used by different studies. Few studies used a high sensitivity measure making under-reporting of levels of contamination a concern. Despite the variable methodology and broadly low sensitivity of the methods used, all activities and all studies identified contamination from droplets that had either settled (splatter) or droplet nuclei remaining in the air as aerosol and at the furthest points studied.

Contamination levels varied with some activities such as hand scaling generating contamination no greater than occurs during speaking [21]. The greatest levels of contamination were found with procedures involving powered devices and water (HSAR and USS). Devices that used air and water together also generated splatter and aerosol and highest nearest the patient. It was not possible to draw conclusions around the use of the slow-speed handpiece with any certainty because no study compared it with anything else for either use of carious tissue removal or orthodontic cement removal.

Although dental procedures are commonly categorised dichotomously as either aerosol producing or non-aerosol producing, this is an over-simplification, bearing in mind that while droplet nuclei ≤ 5 µm in diameter are categorised as aerosols, in reality droplet size lies on a continuum. Since SARS-CoV2 transmission has been reported up to 4 m from the source [43], aerosol transmission remains a possibility.

The majority of studies used a settle plate methodology which is limited to capturing droplets which can carry viruses. Settle plates can detect droplets that have fallen onto a surface. However, air turbulence caused by movement in the surgery may affect what is captured. Air samplers, most of which actively sample the air in the room, will therefore detect both aerosol and airborne droplets before they have fallen to ground (n=23 studies). Many of the studies that stated they were detecting aerosols, did not use a methodology that investigated airborne transmission (i.e. droplets <5 µm) such as air samplers. Most studies’ findings related to droplet splatter detected on settle plates. However, both droplets and aerosols can carry viruses although the universal precautions currently in use will provide protection from droplet transmission.

Although we identified 83 studies investigating eight different procedures, it was difficult to draw definitive conclusions about relative droplet and aerosol generation between procedures. Firstly, because diverse methodologies had been used and secondly, because study quality was generally low, especially in relation to sensitivity of measuring contamination. Low sensitivity could result in significant underestimates: for example, saliva cultured under highly sensitive conditions will yield c 10 CFU/ml but only 10 -10 using low-sensitivity methods. Thirdly, all studies found contamination as far as they measured it and most, for as long as they measured it – which begs the question as to whether greater distances and times measured would have resulted in further positive findings. There was also insufficient data to explore differences according to environmental context such as ventilation or single/multi-chair surgeries (although where multi-surgery environments were studied this showed contamination over larger distances [30, 44].

There has been growing research in this area with over half of the studies being published in the last decade. Most studies (n=62) used microbes purported to be from the oral cavity to monitor contamination from procedures. All, apart from two looking at the bloodborne virus Hepatitis B [45, 46] and three at fungi, investigated bacteria. None of the studies investigated contamination by respiratory viruses. This is despite several significant outbreaks of respiratory viruses where AGPs might have been a risk factor (Severe Acute Respiratory Syndrome (SARS) 2003; Swine flu 2009; Middle East Respiratory Syndrome in 2012). Viruses are difficult to culture, in comparison to bacteria, which may account for this. Most of the studies looked at easily cultured oral bacteria as surrogates of contamination from the droplets and aerosol generated from the procedure. However, viruses are small (typically between 20-300 nm in diameter) and can be carried in the same way as bacteria, therefore patterns of bacterial splatter and aerosol can provide some information to inform viral spread.

None of the studies directly explored exposure (for the dental team or patient) to potentially pathogenic micro-organisms. Studies did, however, identify significant contamination relating to the operator’s head and the patient’s body when powered devices (HSARs and USSs) were used. Visors, glasses and masks were often heavily contaminated, with a small number of ultrasonic studies finding contamination under the visor and mask. The body and operating arm of the operator were subject to significant contamination and studies of the assistant found less contamination (although this varied depending on the area of the mouth being worked on). Areas closest to the patient were most affected. This has implications for decisions about personal protective equipment and the coverage needed. The patient’s face and body were significantly contaminated as a result of powered devices (HSARs and USSs) and were often measured in the oral surgery procedures were also found to be contaminated. No studies investigated contamination of other areas, notably the patient’s (and dentist’s) leg areas but there is no reason to expect them to not be contaminated, given the spread noted across surgery areas in front of the patient. This has implications for infection control measures.

Beyond time, distances and settings there were further obvious gaps in the data as a whole. These include a lack of negative controls and baseline measures, clarity of reporting over specific procedure times and time periods during and following them to see when there was no longer contamination from the procedure. However, the most concerning gap may be the general failure across the studies to report the limits of contamination for distance and very few reporting on time for settle. Studies on mitigating interventions (such as high-volume evacuators, HEPA filters, air changes etc.) may be able to clarify this area further. However, it is encouraging to see more research in the field being undertaken, with three papers published since this search was undertaken, fitting the inclusion criteria [47-49] and one further paper [50]awaiting full text assessment at the time of submission, hopefully these will add to the body of well conducted research informing risk and risk mitigation in relation to AGPs.

This review has focused on droplet and aerosol contamination relating to specific dental procedures, extending beyond previous work that looked at micro-organisms and hazards generated in dental practice [51]. This provides evidence which may be used to determine the baseline risks associated with dental procedures, helping dental professionals to identify clinical situations with increased aerosol transmission risks where mitigation would be strongly advisable.

## Conclusion

Despite generally low sensitivity measures being used, there was evidence of contamination of surfaces around the surgery environment/ personnel or contamination in air from all procedures that were looked at: USS (n=44); HSAR (n=31 studies); oral surgery (n=11), slow-speed handpiece (n=4); air-water (triple) syringe (n=4), air polishing (n=4) hand scaling (n=2), prophylaxis with cup and pumice (n=2). There was evidence that this varied by procedure type. Most studies used microbial surrogate measures (mainly oral microbiota) and blood or colored water for detecting contamination following these procedures. None looked at respiratory viruses. The variability in methodology and variety of outcome measures thwarted attempts to synthesise across studies. By looking at comparisons of procedures within studies, blunt generalisations could be made over higher, moderate and lower risk procedures. There are significant gaps in the evidence and its quality that limit conclusions around all aspects of contamination for different procedures. These hamper evidence-based clinical recommendations and policy decision making, especially relevant for dentistry and COVID-19.

## Data Availability

The supplementary files contain the data files.

## Acknowledgments

We would like to thank library team at the British Dental Journal, for their quick response and dedication to try to supply the full text of papers we required where there was limited access to hard copies in the face of challenges imposed by the COVID-19 lockdown. A special thanks to the American Journal of Dentistry, who provided us a copy of one of their articles at no charge.

## Authors contributions

NI, IJ, RH, JG had oversight of the study planning and execution, and to the conception, design, data acquisition, synthesis, visualisation and interpretation, drafted and critically revised the manuscript. WW conception, design, data synthesis and interpretation, drafted and critically revised the manuscript WA, MR data acquisition, synthesis, visualisation and interpretation, drafted and critically revised the manuscript. SKC, RH, RJ data acquisition, synthesis and interpretation, drafted and critically revised the manuscript. SM design of the study, provision of and management of study literature resources and critically revised the manuscript.

All authors gave their final approval and agree to be accountable for all aspects of the work.

